# Effects of Virtual Co-working Environment Using a Work-Support Application on Subjective Workload, Mood, and Fatigue: A Randomized Crossover Trial

**DOI:** 10.64898/2026.09.19.26362890

**Authors:** Ryo Momosaki, Yoko Hasegawa, Kenta Ushida, Miho Shimizu

## Abstract

**Background:** Digitalization and remote work have increased interest in work environments that reduce workload, negative mood, and fatigue. Low-interaction co-presence in virtual spaces may offer a new digital work environment, but its effects on subjective workload remain unclear.

**Objective:** To examine the effects of the 3D-avatar work-support application gogh on subjective workload, mood, and fatigue. Methods: We conducted an open-label randomized two-period crossover trial in 40 gogh desktop users performing desk- or computer-based tasks. Participants completed 30-min task periods with and without gogh, separated by a 30-min break, in randomized order. The NASA Task Load Index (NASA-TLX) was the primary outcome, with the mood and fatigue domains of the Inventory Scale for Mood and Sense of Fatigue as secondary outcomes. Linear mixed-effects models used condition and period as fixed effects and participant as a random intercept.

**Results:** All 40 participants completed both conditions. NASA-TLX scores were 42.3 ± 12.9 with gogh and 51.7 ± 15.1 without gogh; the adjusted mean difference was −9.5 (95% confidence interval (CI) −13.4 to −5.5). Adjusted mean differences were −11.4 for mood (95% CI −16.6 to −6.2) and −15.0 for fatigue (95% CI −20.3 to −9.7). Exploratory analyses showed lower Frustration, Effort, and Physical Demand with gogh.

**Conclusions:** The gogh condition was associated with a decrease in post-task perceived workload, negative mood, and fatigue. This combined virtual co-working environment may reduce task-related psychological burden, although objective performance, productivity, and long-term occupational health effects require further study.

**Trial registration:** Japan Registry of Clinical Trials (jRCT): jRCT1040260086 https://jrct.mhlw.go.jp/en-latest-detail/jRCT1040260086

## 1. Introduction

In today’s work environments, workload caused by high work intensity, tight deadlines, and extended working hours is a significant occupational health issue that can impact workers’ health, long-term engagement, and performance. A survey of 25 European OECD countries indicated that, in 2021, 13% of workers experienced job strain, which occurred when job demands surpassed available resources, and 73% were exposed to work intensity such as fast work tempo and tight deadlines [1]. Psychosocial risks, such as excessive workload, can have an impact not just on stress and exhaustion but also on physical and mental health, as well as job performance [2]. As a result, developing work settings that effectively regulate workload, mood, and fatigue is critical for promoting sustained work and steady work performance without jeopardizing health.

Sensory cues such as music and the presence of others have been used to influence the work environment. Music has been shown to affect cognitive task performance and subjective workload [3,4]. Coaction, which involves individuals performing autonomous work in the same environment as others, is a type of social facilitation. Claypoole et al. found that the presence of an independent coactor improved performance on a sustained-attention task while reducing subjective burden [5]. However, social facilitation effects are not consistent and may vary depending on job difficulty, the other person’s function, and how others’ social presence is perceived [6–8].

Recent technologies allow us to depict the presence of others in virtual rather than real spaces. In virtual reality (VR) and collaborative virtual environments, avatars can foster a sense of social presence and co-presence with others in the same space [9,10]. These perceptions differ depending on whether the avatar is human-controlled and how much visual and nonverbal cues are provided [9,11,12]. In shared virtual worlds, nonverbal information, such as gaze, might influence interpersonal perception and task performance [13]. Recent studies have examined how avatar fidelity and avatar-mediated communication affect co-presence, attention allocation, and perceptions of social interaction [14,15]. Studies have also measured workload, presence, and emotion recognition in collaborative virtual environments [16], as well as the effect of avatars on peer social presence [17]. Thus, the presence of virtual persons, such as coactors in real environments, may influence workers’ psychological moods and task experiences.

However, in terms of office design, a significant disparity remains. The majority of studies on social facilitation in VR have used preset experimental tasks such as games, training, collaborative manipulation, or education. For example, research on remote collaboration has investigated situations in which participants work together to complete a common job [18]. In contrast, nothing is known about the impact of low-interaction co-presence, in which “other people are also doing their own work” without active engagement, on perceived workload, mood, and fatigue during ordinary desk work. A digital work environment that provides social co-presence without increasing interpersonal coordination, particularly for individual work in home or remote settings, may be worth investigating as a type of work design important to both occupational health and productivity.

gogh is a 3D-avatar work-support application that encourages concentration. As of July 2026, the mobile version had received over 3 million cumulative downloads and was frequently utilized [19]. In the multiplayer mode, multiple users enter the same virtual world and operate independently. Because active conversation or a shared task is not required, this environment differs from traditional collaborative VR and can be described as a virtual co-working setting in which users “do their own work while sensing the presence of others,” similar to a library, study room, or co-working space. If such an environment reduces workload and fatigue, it could provide a location-independent digital work environment that promotes worker well-being and long-term task performance. However, its impacts have not been thoroughly investigated.

Using a randomized crossover design, this study compared a combined virtual co-working condition in which participants utilized gogh to work in the same virtual space as other users’ avatars to a usual-work control condition without gogh, and investigated the effect on subjective workload. We also assessed the impact on mood and fatigue. The study sought to evaluate whether low-interaction avatar-mediated co-presence may reduce workload while also providing foundational data for occupational health, digital workplace design, and future evaluation of productivity.

## 2. Methods

### 2.1 Study design

This was an open-label randomized two-period crossover trial that compared a condition in which participants used the work assistance application gogh to work in the same virtual space as other users’ avatars (the gogh virtual co-working condition) to a usual-work condition without gogh. Participants were registered between July 24 and August 31, 2026. The study was approved by the Medical Research Ethics Review Committee of Mie University Hospital (approval no. H2026-164) and prospectively registered in the Japan Registry of Clinical Trials (jRCT1040260086).

### 2.2 Participants

Eligible participants were required to (1) be at least 18 years old; (2) typically undertake desk or computer-based tasks such as writing or illustration; (3) utilize the desktop version of gogh; and (4) understand the study and provide voluntary consent to participate. Exclusion criteria included a physical or mental condition that made continuous work performance difficult, an inability to secure a reliable internet connection, or trouble logging into or using the gogh desktop application’s multi-room function.

Participants were solicited online using a study-affiliated X account. Google Forms were used to offer study information, and individuals provided consent to participate after reviewing it. After completing the study, participants received an Amazon gift card worth JPY 1,500. Prior to the intervention, Google Forms was used to obtain baseline information, including age, sex, cumulative gogh use time, occupational/social status, and routine activities performed when using gogh. Occupational/social status and routine gogh activities were collected as free-text responses and grouped into broad categories during the analysis. Because several participants indicated more than one routine gogh activity, their results were aggregated into multiple responses.

In a previous study, the global NASA-TLX score during the Attention Network Test was higher under high-arousing music than under silence, with an effect size of Cohen’s d = 0.60 [3]. Because there was no directly comparable impact estimate for virtual co-working, we used Cohen’s d = 0.50 for sample-size planning. Using a two-sided significance level of 5% and 80% power, G*Power 3 was used to calculate the required sample size of 34 [20]. Allowing for attrition, the target sample size was set at 40.

### 2.3. Randomization and intervention

Participants were randomized into one of two intervention sequences. Following enrollment, a system created with Google Forms performed randomization based on the Fisher-Yates shuffle and assigned the intervention sequence. Because allocation was created automatically following participant enrollment, enrollment personnel were unable to predict the allocation of the next participant, resulting in allocation concealment. In one sequence, participants worked for 30 min with gogh, then had a 30-min break before repeating the assignment without gogh. In the alternative sequence, participants first worked for 30 min without gogh, then took a 30-min break before working for 30 min with gogh.

Tasks included participants’ typical tasks, such as writing, drawing, programming, or studying, and were completed under conditions similar to real-life use. To improve within-participant comparability, participants were required to complete tasks that were as identical as possible between the two conditions, with each work period standardized to 30 min. In the gogh condition, participants used the desktop version of gogh (Ver. 3.2.1) and logged into a multi-room with at least two other users present when the task began (Figure 1). The avatar type was not provided. Because music might impair cognitive performance and subjective workload during tasks [3,4], the in-app music/background music feature was disabled, and music playback from external devices was disallowed. Ambient sounds within gogh and sounds generated with avatars were not prohibited. A 30-min break was allowed between conditions. Short washout periods of about 30 min have also been utilized in randomized crossover trials with comparable mental workloads [4]. The tasks performed during the trial were recorded as free-text responses.

**Figure 1.**
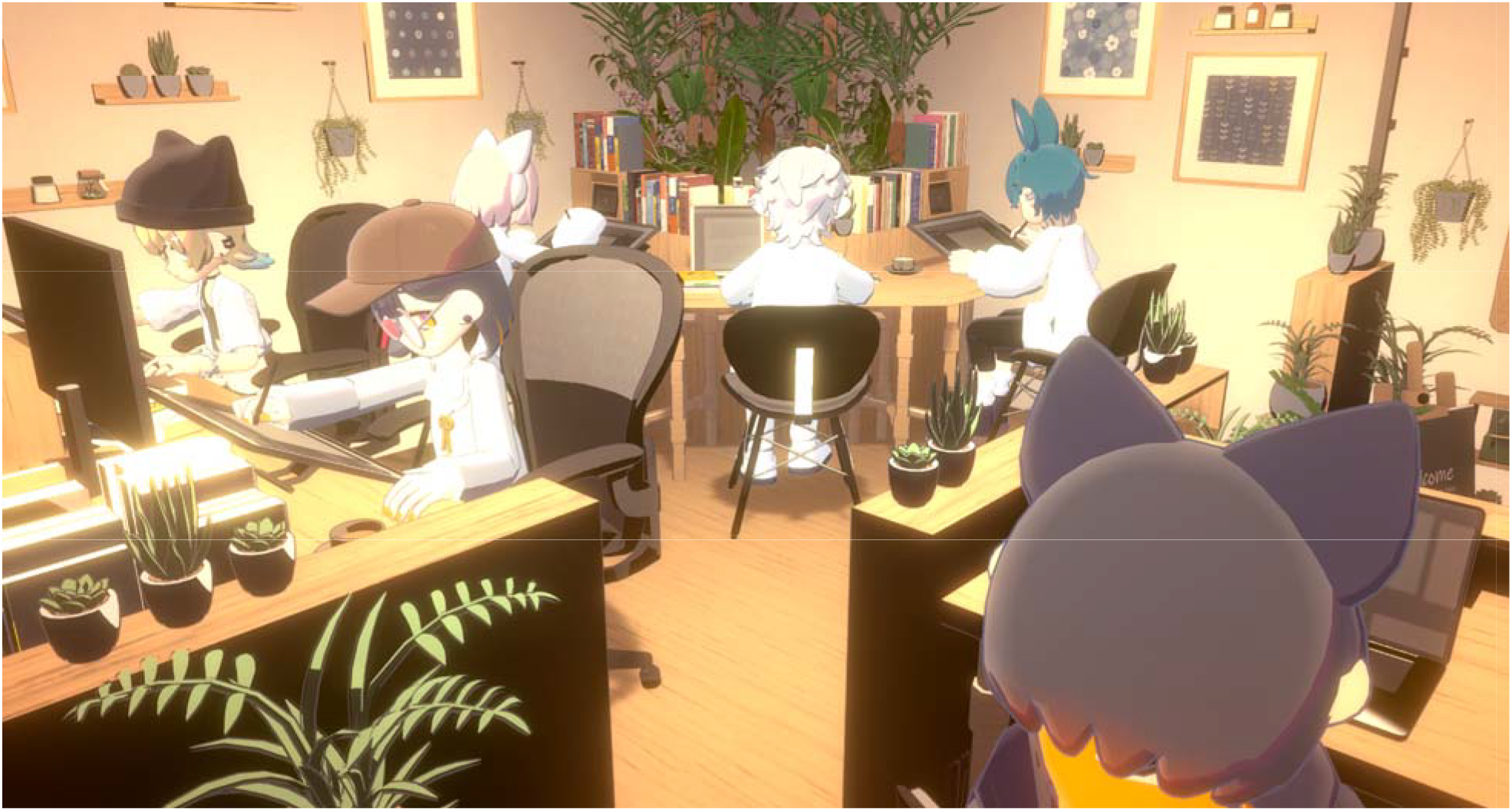
Example of the gogh virtual co-working environment. Multiple users’ avatars occupy the same virtual room while independently performing their own tasks.

### 2.4 Outcome measures

The primary outcome was subjective workload assessment using the NASA Task Load Index (NASA-TLX). Secondary outcomes included the six NASA-TLX subscales, as well as the mood state and fatigue domains of the Inventory Scale for Mood and Sense of Fatigue (SMSF). Outcomes were collected using Google Forms, and participants completed the NASA-TLX and SMSF immediately after each 30-min work period. Following each condition, an optional free-text field area was also provided to obtain feedback on participants’ subjective experiences with the task.

The NASA-TLX is a subjective workload measure that includes six domains: Mental Demand, Physical Demand, Temporal Demand, Performance, Effort, and Frustration [21,22]. Each item was graded on an 11-point scale from 0 to 10 and then linearly translated into a 0-100 scale by multiplying by 10. For the Performance item, higher scores indicated poorer perceived performance. The mean of the six items was calculated as the raw NASA-TLX, with lower scores indicating lower subjective workload.

The SMSF measures subjective mood and fatigue, and it has been shown to be reliable and valid [23]. This study employed six items to assess mood state and four items to assess fatigue. Each item was scored from 0 to 10, multiplied by 10 to convert the score to a 0-100 scale, and the average was calculated within each domain. Lower SMSF scores suggest less negative mood and fatigue.

### 2.5 Statistical analysis

Continuous variables are presented as means and standard deviations, while categorical variables are presented as counts and percentages. Because cumulative gogh use time was expected to be skewed, it is represented by the median and interquartile range. Crossover studies must take into account both the intervention condition and the time period, as well as any within-participant correlation [24]. In the primary analysis, linear mixed-effects models (LMMs) were used with each outcome as the dependent variable, intervention condition (gogh/control) and time (first/second) as fixed effects, and participant as a random intercept. The intervention effects are shown as adjusted mean differences computed as gogh minus control, with 95% confidence intervals (CIs). The same model was used for the six NASA-TLX subscales. The primary outcome tests were two-sided, with P < 0.05 indicating statistical significance. NASA-TLX subscale analyses ere exploratory, with no adjustment for multiple comparisons. Statistical analyses were carried out with Python version 3.13.5 and statsmodels version 0.14.6, with LMMs computed using maximum likelihood.

### 3. Results

### 3.1 Participants

During the study period, 40 individuals were enrolled, and all of them completed both conditions and replied to the postcondition assessments; therefore, all 40 were included in the analysis. Twenty-six participants (65.0%) were aged 18–29 years, 10 (25.0%) were aged 30–39 years, and 4 (10.0%) were aged 40–49 years (Table 1). Thirty-two participants (80.0%) were female, 7 (17.5%) were male, and 1 (2.5%) selected “Other.” Median cumulative gogh use was 165 h (interquartile range, 30.8– 463.9 h). Twenty-two participants (55.0%) were assigned to the gogh-first sequence and 18 (45.0%) to the control-first sequence (Figure 2). The most prevalent occupational/social category was creator or creative professional (17 participants, 42.5%), followed by student (9, 22.5%) and company employee/professional (8, 20.0%). The most common routine work activity was illustration, manga, or design (22 participants, 55.0%), followed by writing or text creation (14 participants, 35.0%). During the experiment, tasks included creative work such as drawing, manga, or design, as well as writing, study/research, video editing, programming, and work-related document preparation. Tasks in the first and second periods were largely identical or similar among participants. At the start of the gogh condition, the multi-room had a median number of 6 users, including the participant (interquartile range, 3.0–8.3). Participants employed a range of multi-rooms; some were designed for creative or focused work, while others resembled cafes, lounges, or private rooms.

**Table 1.** Participant characteristics.

| Item | Overall (N = 40) |
| --- | --- |
| Age 18–29 years | 26 (65.0%) |
| Age 30–39 years | 10 (25.0%) |
| Age 40–49 years | 4 (10.0%) |
| Female | 32 (80.0%) |
| Male | 7 (17.5%) |
| Other | 1 (2.5%) |
| Cumulative gogh use, median (interquartile range), h | 165 (30.8–463.9) |
| gogh-first sequence | 22 (55.0%) |
| Control-first sequence | 18 (45.0%) |
| Occupational/social status |  |
| Creator/creative professional | 17 (42.5%) |
| Student | 9 (22.5%) |
| Company employee/professional | 8 (20.0%) |
| Unemployed/homemaker | 4 (10.0%) |
| Other | 2 (5.0%) |
| Routine activities performed while using gogh (multiple responses) |  |
| Illustration/manga/design | 22 (55.0%) |
| Writing/text creation | 14 (35.0%) |
| Study/research/information gathering | 7 (17.5%) |
| Work/administrative/document preparation | 7 (17.5%) |
| Video editing/streaming | 7 (17.5%) |
| Programming/3D/Unity | 6 (15.0%) |

Note: Routine activities conducted while using gogh were summarized as multiple responses; therefore, percentages do not equal 100%.

**Figure 2.**
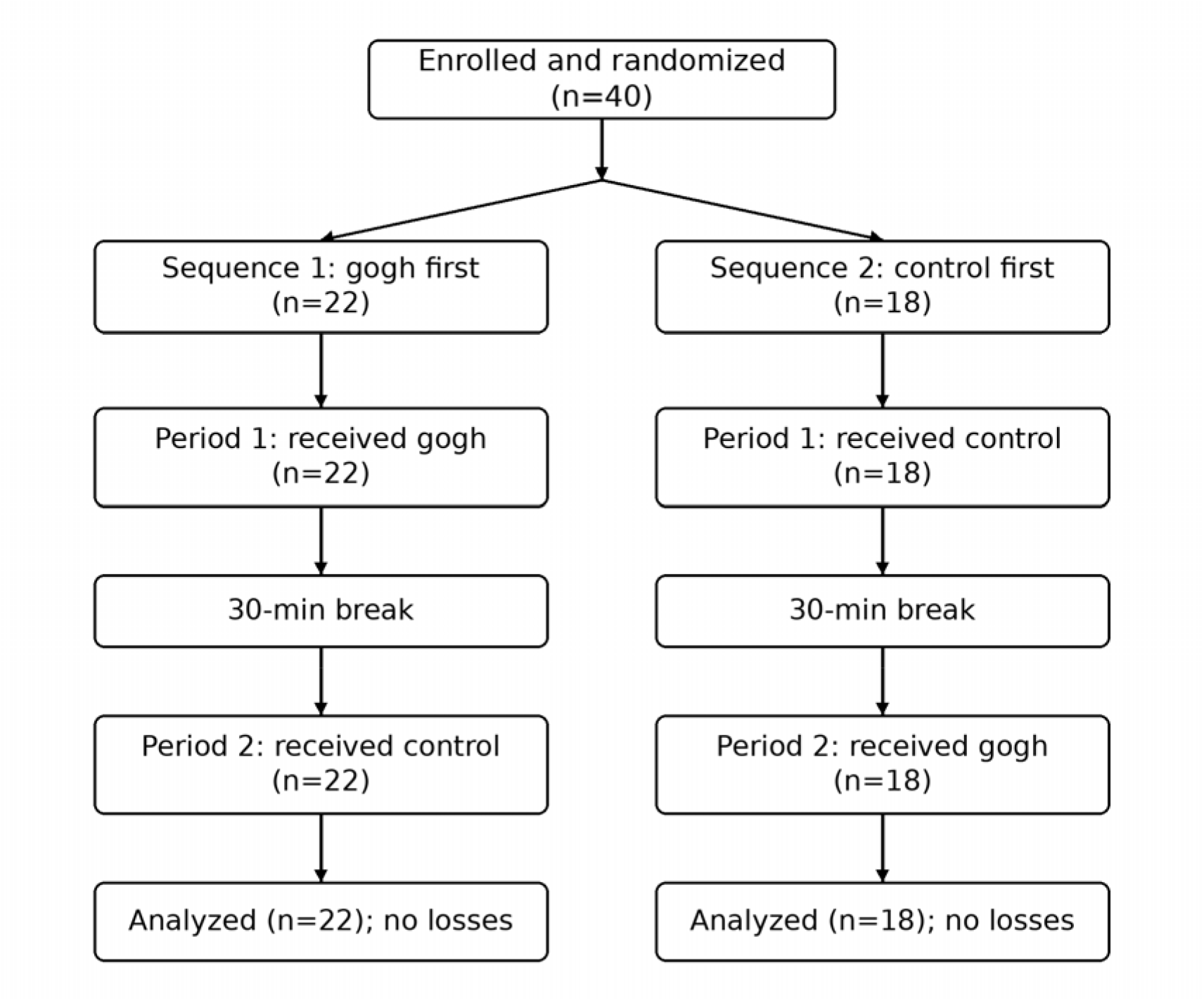
Participant flow through the randomized two-period crossover trial.

### 3.2 Primary outcome: NASA-TLX

Following the task, the mean NASA-TLX scores were 42.3 ± 12.9 in the gogh condition and 51.7 ± 15.1 in the control condition. The adjusted mean difference from the LMM (gogh minus control) was −9.5 points (95% CI −13.4 to −5.5; P < 0.001). The period effect was not statistically significant (P = 0.475) (Table 2).

**Table 2.** Comparison of primary and secondary outcomes.

| Outcome | gogh<br>Mean ± SD | Control<br>Mean ± SD | Adjusted mean<br>difference | 95% CI | P value | P-value for<br>period effect |
| --- | --- | --- | --- | --- | --- | --- |
| NASA-TLX | $42.3 \pm 12.9$ | $51.7 \pm 15.1$ | $-9.5$ | $-13.4$ to $-5.5$ | $<0.001$ | 0.475 |
| SMSF mood<br>state | $22.7 \pm 15.5$ | $33.7 \pm 23.9$ | $-11.4$ | $-16.6$ to $-6.2$ | $<0.001$ | 0.102 |
| SMSF fatigue | $36.0 \pm 21.3$ | $50.5 \pm 22.8$ | $-15.0$ | $-20.3$ to $-9.7$ | $<0.001$ | 0.073 |

Adjusted mean differences were calculated as gogh minus control. CI, confidence interval; SD, standard deviation

### 3.3 SMSF

Immediately after the task, the mean SMSF mood state scores were 22.7 ± 15.5 in the gogh condition and 33.7 ± 23.9 in the control condition, with an adjusted mean difference of −11.4 points (95% CI −16.6 to −6.2; P < 0.001). Mean SMSF fatigue scores were 36.0 ± 21.3 and 50.5 ± 22.8, respectively, with an adjusted mean difference of −15.0 points (95% CI −20.3 to −9.7; P < 0.001). There were no statistically significant period effects on mood state (P = 0.102) or fatigue (P = 0.073) (Table 2).

### 3.4 NASA-TLX subscales

The six NASA-TLX subscales were investigated with the same LMM as in the primary analysis (Table 3). Compared to the control condition, the gogh virtual co-working condition exhibited lower Frustration (adjusted mean difference −22.2, 95% CI −31.1 to −13.3; P < 0.001), Effort (−16.2, 95% CI −24.1 to −8.4; P < 0.001), and Physical Demand (−10.9, 95% CI −17.8 to −4.0; P = 0.002). There were no significant changes across conditions in terms of Mental Demand, Temporal Demand, or Performance.

**Table 3.** Comparison of NASA-TLX subscales.

| Subscale | gogh<br>Mean ± SD | Control<br>Mean ± SD | Adjusted mean<br>difference | 95% CI | P value | P-value for<br>period effect |
| --- | --- | --- | --- | --- | --- | --- |
| Mental Demand | $67.5 \pm 22.3$ | $67.3 \pm 25.3$ | $-0.3$ | $-6.7$ to $6.1$ | 0.920 | 0.077 |
| Physical Demand | $33.3 \pm 25.1$ | $44.8 \pm 28.6$ | $-10.9$ | $-17.8$ to $-4.0$ | 0.002 | 0.092 |
| Temporal Demand | $42.3 \pm 26.1$ | $44.5 \pm 27.4$ | $-2.7$ | $-13.1$ to $7.8$ | 0.618 | 0.451 |
| Performance | $38.0 \pm 25.8$ | $43.3 \pm 22.9$ | $-4.6$ | $-13.0$ to $3.8$ | 0.285 | 0.112 |
| Effort | $44.3 \pm 25.0$ | $60.0 \pm 23.0$ | $-16.2$ | $-24.1$ to $-8.4$ | $<0.001$ | 0.226 |
| Frustration | $28.8 \pm 24.6$ | $50.3 \pm 30.2$ | $-22.2$ | $-31.1$ to $-13.3$ | $<0.001$ | 0.139 |

Adjusted mean differences were calculated as gogh minus control. All analyses were exploratory, and no adjustment for multiple comparisons was applied. CI, confidence interval

In the free-text responses, participants in the control condition stated that they were “easily distracted,” “bored,” or found the lack of background music or ambient sound “uncomfortable.” Participants in the gogh condition reported feeling calmer as a result of working with others and experiencing mood changes as the scenery in the virtual space changed.

## 4. Discussion

In this study, NASA-TLX scores were lower in the gogh condition, in which participants worked independently in the same virtual space as other users represented by 3D avatars, than in the control condition. The SMSF also revealed fewer levels of negative mood state and fatigue. Frustration, Effort, and Physical Demand were lower on the NASA-TLX subscales, but there were no significant differences in Mental Demand, Temporal Demand, or Performance. These findings imply that gogh’s unified digital work environment may lower subjective costs such as perceived effort and discomfort while doing tasks.

The reduction in NASA-TLX found in this study is in line with coactor research in physical environments. In a vigilance task involving 100 participants, Claypoole et al. reported fewer false alarms and lower subjective workload when an independent coactor was present than when participants performed the task alone [5]. Research in social facilitation has also demonstrated that impacts on performance, workload, and stress differ depending on the role of others, such as a supervisor, and the context of electronic monitoring [25–28]. Taken together, the presence of others may be more than just a source of distraction; it may modify task engagement, arousal, and self-regulation, resulting in a shift in subjective workload. Because similar patterns were observed in the present study despite the absence of physically co-present others, the workload reduction related to coaction may apply to avatar-mediated environments.

However, just inserting avatars of other people in a virtual space does not always result in uniform reductions in workload. Avatar agency and anthropomorphism have been shown to affect perceptions of telepresence, co-presence, and social presence [9]. A meta-analysis also found that social presence is higher when a virtual human is seen as an avatar directed by a human rather than a computer-controlled agent [29]. Furthermore, avatar fidelity affects co-presence [14], and nonverbal information conveyed by avatars is linked to attention allocation and perceptions of social interaction [15]. These findings imply that the influence of others in virtual settings may be dependent not just on the presence of an avatar but also on how the person behind that avatar is viewed.

In gogh, avatars in the same room are controlled by real users, with each working on their own task at the same time. The perception of “working in parallel in the same place with real other people” could have altered participants’ psychological experiences during the task, contributing to the observed effort reduction. Parallel work with real others may lessen psychological stress, which is also consistent with the observed reductions in Frustration and Effort in this study.

Others’ influence should not be confused with making the task simpler. In this study, mental and temporal demands changed little; therefore, it is doubtful that the tasks completed or their time limits were easier. Rather, the sense of participating in action with others may have aided task engagement and persistence. According to research on virtual co-working, ICT-based virtual co-working spaces can promote social proximity and motivation [30]. The use of VR/AR in co-working spaces has also been advocated as a way to promote location-independent social interaction and collaboration [31]. Thus, gogh’s effect may be less about “making work easier” and more about “reducing the psychological cost of continuing to work.” This could provide a work-design advantage, especially for individual desk work and remote work, by improving the work environment while reducing interruptions or interpersonal coordination costs.

The SMSF results support the NASA-TLX findings by improving both mood and fatigue. Workload is linked to both cognitive demands and subjective experiences such as effort, stress, and fatigue. Studies on social facilitation have revealed that performance increases in the presence of others do not always modify workload or stress in the same direction [27,28], showing that psychological responses vary depending on the role of others and the context. In the gogh environment, the other users were not supervisors or evaluators but people conducting their own work, much like the participants. This non-evaluative parallel co-working connection may have given social presence without increasing emotions of monitoring or evaluation apprehension, resulting in improved mood and fatigue. An essential next step for occupational health is to investigate whether such reductions in psychological burden are associated with fatigue accumulation and work engagement with prolonged or repeated use.

When examining applications for job participation and occupational rehabilitation, it is critical to distinguish between “directly improving work capacity” and “creating an environment that makes it easier to continue engaging in work.” In a 2025 VR rehabilitation trial in persons with chronic stroke, the addition of a virtual agent appeared to improve task engagement and tenacity in training, but its effect on game performance was limited [32]. This tendency is congruent with the present findings, where changes were noticed more clearly in subjective experiences such as Effort, Frustration, and fatigue than in Mental Demand or Performance. The presence of virtual others may therefore have an impact on ease of engagement, persistence, and psychological burden rather than directly enhancing capacity or productivity. Future studies could investigate this approach in contexts requiring sustained work involvement, such as remote work for general workers, returnto-work support, and occupational rehabilitation. However, the present sample was not representative of the overall workforce and did not include participants receiving return-to-work support; therefore, these prospective applications remain hypothetical.

Conversely, more social presence in collaborative VR may not necessarily result in lower workload. In collaborative virtual environments, workload and presence have been shown to change by task and across time [16]. In a remote collaboration system, visual signals such as avatars and gestures boosted social presence and co-presence, but the NASA-TLX workload assessment showed no significant difference [33]. A meta-analysis of virtual-character agency discovered that avatars had a stronger social presence than agents, but the effects on behavioral outcomes were unclear [29]. One possible distinction from the present study is that gogh does not require users to undertake a shared task, which may reduce the additional cognitive strain involved with coordination and communication. In other words, its “low-interaction” design may minimize workload by limiting interaction costs while preserving the potential benefits of social presence.

The observed variations cannot be attributed to a single component of gogh. The gogh condition consisted of several aspects, including the presence of other users’ avatars, the participant’s own avatar, the 3D environment, and ambient sounds. Because the control condition did not include matched visual or ambient-sound stimulation, the observed effects cannot be attributed specifically to avatar-mediated copresence. We also did not directly assess social presence, co-presence, motivation, loneliness, or task engagement. As a result, a mediating mechanism that reduces workload through enhanced co-presence remains hypothetical.

This study has several limitations. First, participants were previous gogh users, and users with relatively positive prior experiences may have been more willing to participate; thus, caution should be exercised when generalizing the findings to inexperienced users or the general population. The sample was also disproportionately young and female, with many participants working in artistic fields, which limits its applicability to larger working populations. Second, the nature of the intervention prevented participants from being blinded, and anticipation effects may have influenced the self-reported outcomes. Third, tasks were not totally standardized and instead reflected participants’ typical activities; variations in task content may have influenced the outcomes. In contrast, this allowed for examination under more realistic settings. Fourth, the interval between conditions was 30 min, so the effects of the previous condition may not have been eliminated. Although period was included in the model, a nonsignificant period effect does not imply the absence of carryover effects. Fifth, results were limited to self-report measures collected immediately following each 30-min task. Objective indicators such as work quantity, accuracy, and productivity, as well as consequences of repeated or long-term use, were not assessed. Sixth, because social presence and copresence were not quantified, the psychological mechanisms underlying the observed differences could not be explicitly tested.

Overall, this study broadens the scope of social facilitation and workload reduction described in physical coaction research to avatar-mediated virtual co-working throughout daily tasks. The consistent reductions in post-task subjective workload, negative mood, and fatigue in an environment that did not require direct collaboration or communication have implications for the design of digital work environments. However, productivity was not directly tested; therefore, it is unclear whether reduced burden leads to improvements in work quantity, accuracy, efficiency, or long-term work performance. Future research should include inexperienced users as well as more diverse working populations, evaluating objective task performance, social presence, work engagement, fatigue accumulation, and long-term work participation.

## 5. Conclusions

In the combined virtual co-working condition, participants worked for 30 min in the same virtual space as others via 3D avatars utilizing gogh and reported reduced post-task subjective workload, negative mood state, and fatigue compared with the usual-work control condition. Exploratory analyses of NASA-TLX subscales revealed decreased levels of Frustration, Effort, and Physical Demand. These findings suggest that the gogh-based combined virtual co-working condition may alleviate task-related psychological burden in digital work environments. However, the effects on objective productivity and long-term occupational health were not assessed, and additional research is required to clarify these outcomes and the contributions of particular intervention components.

## Preprint notice

This manuscript is a preprint and has not been certified by peer review.

## Ethics approval and consent to participate

This study was conducted in accordance with the Declaration of Helsinki and the Ethical Guidelines for Medical and Biological Research Involving Human Subjects and was approved by the Medical Research Ethics Review Committee of Mie University Hospital (approval no. H2026-164). Electronic informed consent to participate was obtained from all participants.

## Trial registration

Japan Registry of Clinical Trials (jRCT), jRCT1040260086. Registered prospectively.

## Data availability

The anonymized data used in this study are available from the corresponding author upon reasonable request, within the limits of ethical and legal restrictions and participant consent.

## Author contributions

Ryo Momosaki: study conception, study design, study supervision, data interpretation, and manuscript preparation. Yoko Hasegawa: study conduct, data collection, and manuscript review. Kenta Ushida: statistical analysis, data interpretation, and manuscript review. Miho Shimizu: data management, data quality control, and manuscript review. All authors reviewed and approved the manuscript and consented to its posting as a preprint.

